# Clofazimine population pharmacokinetics and target attainment in rifampicin-resistant Tuberculosis patients

**DOI:** 10.64898/2025.12.04.25340990

**Authors:** Bern-Thomas Nyang’wa, Ilaria Motta, Ronelle Moodliar, Varvara Solodovnikova, Shakira Rajaram, Mohammed Rasool, Catherine Berry, Zhonghui Huang, Geraint Davies, David Moore, Frank Kloprogge

## Abstract

Clofazimine is a key component of the BPaLC regimen (bedaquiline, pretomanid, linezolid, and clofazimine), an investigational treatment for rifampicin-resistant tuberculosis (RR-TB). However, clofazimine’s elimination half-life has never been formally characterized amongst tuberculosis patients and a pharmacokinetic (PK) - pharmacodynamic (PD) endpoint for probability of target attainment (PTA) evaluations is lacking. To support dose optimization and efficacy interpretation, we developed a clofazimine population pharmacokinetic (PK) model and evaluated exposure and PTA. Thirty RR-TB patients received daily oral clofazimine at 100 mg, and plasma samples were collected at multiple time points that covered clofazimine wash-out after treatment completion. Clofazimine concentrations were quantified using high-performance liquid chromatography-tandem mass spectrometry and PK modeling was performed using nlmixr2 in R. A two-compartment model with first-order absorption and elimination, and body weight allometric scaling best described the data. Typical clearance was 6.84 L/h, median AUC₀₋₂₄ was 1.44 mg·h/L and 15.67 mg·h/L at week one and end of treatment, respectively; and median trough concentration was 0 mg/L and 0.66 mg/L at baseline and end of treatment. Simulations using 100mg daily dose showed ≥90% T>MIC during a dose interval, provided MIC’s are 0.5 mg/L or lower. For the TB-PRACTECAL *Mtb* isolates’ MIC distribution, this translates into over 95% of the trial participants. We reported clofazimine’s post-treatment population PK characteristics and confirmed clofazimine’s long terminal half-life of 45 days. However, robust clinical PK-PD targets for PTA when given as combination treatment remain unavailable and thus further investigation is warranted.

## Introduction

Tuberculosis is one of the top ten causes of death and the leading cause of death from a single infectious agent, responsible for approximately 1.23 million fatalities in 2023. Rifampicin-resistant TB remains a public health threat(1). Clofazimine is a group B anti-TB drug and included in several new RR-TB treatment regimens, including the Bedaquiline, pretomanid, linezolid, and clofazimine (BPaLC) has been trialed for treatment of rifampicin-resistant TB,in the TB-PRACTECAL trial (2).

Clofazimine is a lipophilic riminophenazine licensed for treatment of leprosy. Several mechanisms of action have been postulated which may predominate depending on the specific physiological environment, some of these include intracellular redox cycling, interfering with potassium uptake in membrane phospholipids and anti-inflammatory activity through inhibition of T-lymphocytes activation and proliferation(3).

Oral administration of clofazimine 100mg daily in leprosy patients results in average plasma levels of 0.7 mg/L. High fat food increases bioavailability by 45%. When dosed at 300mg for the first three days and then 100mg for the remaining 11 days, the Cmin and Cmax at day 14 are 0.153mg/L and 0.232mg/L respectively.

Clofazimine is partially metabolised in the liver, but the full scope of its metabolic pathways is not known. Negligible amount of parent drug or metabolites are found in urine; however, a significant amount is found in faeces(4–7).

Clofazimines elimination half-life is long, although its population pharmacokinetic (PK) characteristics amongst tuberculosis patients remain understudied, especially with respect to its elimination clearance since previous studies lack extended plasma sampling beyond last dose intake(6, 8–10). Clofazimine is also highly protein bound, mice model studies have demonstrated the free drug to be less than 15% while others have even suggested it to be lower than 1%(11, 12).

Despite *in-vitro* promise and hypothesised added value in shortening of treatment regimens, demonstrating the direct efficacy of clofazimine in patients with TB has proven elusive and consequently an effective dose and pharmacokinetic-pharmacodynamic (PK-PD) target has not yet been established(4, 13, 14). The duration of serum drug concentration above the MIC has been associated with clofazimine’s sustained antimicrobial activity(15, 16).

This study therefore studied clofazimine’s PK properties amongst rifampicin resistant tuberculosis patients using data covering the drugs terminal elimination phase. The study also evaluated the PK-PD endpoint against prospectively collected *Mtb* susceptibility data(2).

## Results

A total of 30 study participants (23% female) with a median age of 35 years (range: 19 – 50 years) contributed 286 timed plasma samples which upon bioanalysis were included in the clofazimine population PK dataset (Table 1). A total of 23 samples were collected before the first dose with the remaining 263 samples being collected after the first dose and 38 samples were below the limit of quantification. The observed concentrations of clofazimine ranged from 10.78 – 1467.85 ng/ml. The median trough concentration was 365.40 ng/ml, with an interquartile range of 102.51 – 528.55 ng/ml.

**Table 1:** Baseline characteristics of the study participants.

|  |  |
| --- | --- |
| Sample size (n) | 30 |
| Age (y) | 39 (23-53) |
| Baseline weight (kg) | 56.8 (39.2-104) |
| Body mass index (kg/m<sup>2</sup>) | 19.3 (14.3-28.2) |
| Fat free mass (kg) | 47.1 (28.6-75.5) |
| BUN (mmol/L) | 3.5 (1.8-8.5) |
| ALT (IU/L) | 19 (4-77) |
| AST (IU/L) | 22 (10-58) |
| Total protein (g/L) | 75 (67-107) |
| Estimated creatinine clearance (mL/min) | 104.1 (60.5-175.9) |
| Female, n (%) | 7 (23.3) |
| Caucasian race, n (%) | 12 (40) |
| Black race, n (%) | 17 (56.7) |
| Asian race, n (%) | 1 (3.3) |
| HIV positive, n (%) | 10 (33.3) |

Median (min-max) if not stated otherwise. BUN = blood urea nitrogen, ALT= alanine transaminase, AST= aspartate aminotransferase.

A two-compartment first order absorption and elimination model with a fixed lag time and absorption constant from a previous publication best described the clofazimine PK data(10). Random effects on clearance, central volume of distribution, peripheral volume of distribution and inter-compartmental model were included in the model to explain the inter-individual variability. A combined residual error model was used for the unexplained variability (Table 2 and Appendix 1).

**Table 2:** Estimated clofazimine population pharmacokinetic parameters

| Parameter | Parameter estimate<br>(%RSE) | Bootstrap<br>Median (95% CI) |
| --- | --- | --- |
| **Fixed-effect parameters** |  |  |
| $k_a$ (hr<sup>-1</sup>) | 0.67 (fixed) | 0.67 (fixed) |
| T<sub>lag</sub> (hr) | 0.62 (fixed) | 0.62 (fixed) |
| CL/F (L/hr) | 6.84 (7.49) | 6.91 (5.83 – 8.67) |
| V<sub>c</sub>/F (L) | 1,750 (3.43) | 1,800 (1,640 – 2,430) |
| Q/F (L/hr) | 41.7 (5.67) | 42.3 (37.9 – 56.4) |
| V<sub>p</sub>/F (L) | 9,150 (1.17) | 9,200 (7,730 – 11,000) |
| **Random-effect parameters** |  |  |
| CL/F %CV,<br>[shrinkage %] | 77.3 [8.78] | 74.6 |
| V<sub>c</sub>/F % CV,<br>[shrinkage %] | 173 [2.39] | 173 |
| Q/F % CV,<br>[shrinkage %] | 82 [26.1] | 81.5 |
| V<sub>p</sub>/F % CV,<br>[shrinkage %] | 44.6 [40.9] | 44.5 |
| **Residual error parameters** |  |  |
| Proportional | 0.198 | 0.195 (0.146 – 0.246) |
| Additive<br>(mg/L) | 0.0164 | 0.0164 (0.016 – 0.0165) |

$k_a$ : absorption rate constant, T<sub>lag</sub>: lag time of absorption, CL/F: elimination clearance, V<sub>c</sub>/F: central volume of distribution, Q/F: Inter-compartmental clearance, V<sub>p</sub>/F: peripheral volume of distribution. %RSE = percentage relative standard error. Allometric bodyweight scaling on CL/F and V/F had fixed power coefficients of 0.75 and 1.

Allometric body weight scaling on clearance and volume of distribution was embedded a priori, and blood urea nitrogen ( BUN), creatinine clearance, total protein, aspartate aminotransferase (AST), race, Human immunodeficiency virus (HIV) infection, female sex and treatment regimen were included in the forward first step analysis (p < 0.05), although none of these could be retained in the backward step (p < 0.001) (Appendix 2).

Goodness-of-fit plots (GOF) for the clofazimine population PK model showed no significant bias from the unity line, indicating that the model predicted individual and population values closely matched the observed PK data (Figure 1). Model validation using a visual predictive check (VPC, Figure 2) plot visually confirmed the predictive accuracy of the clofazimine population PK model. Individual drug profiles were included in the Appendix 3.

**Figure 1:**
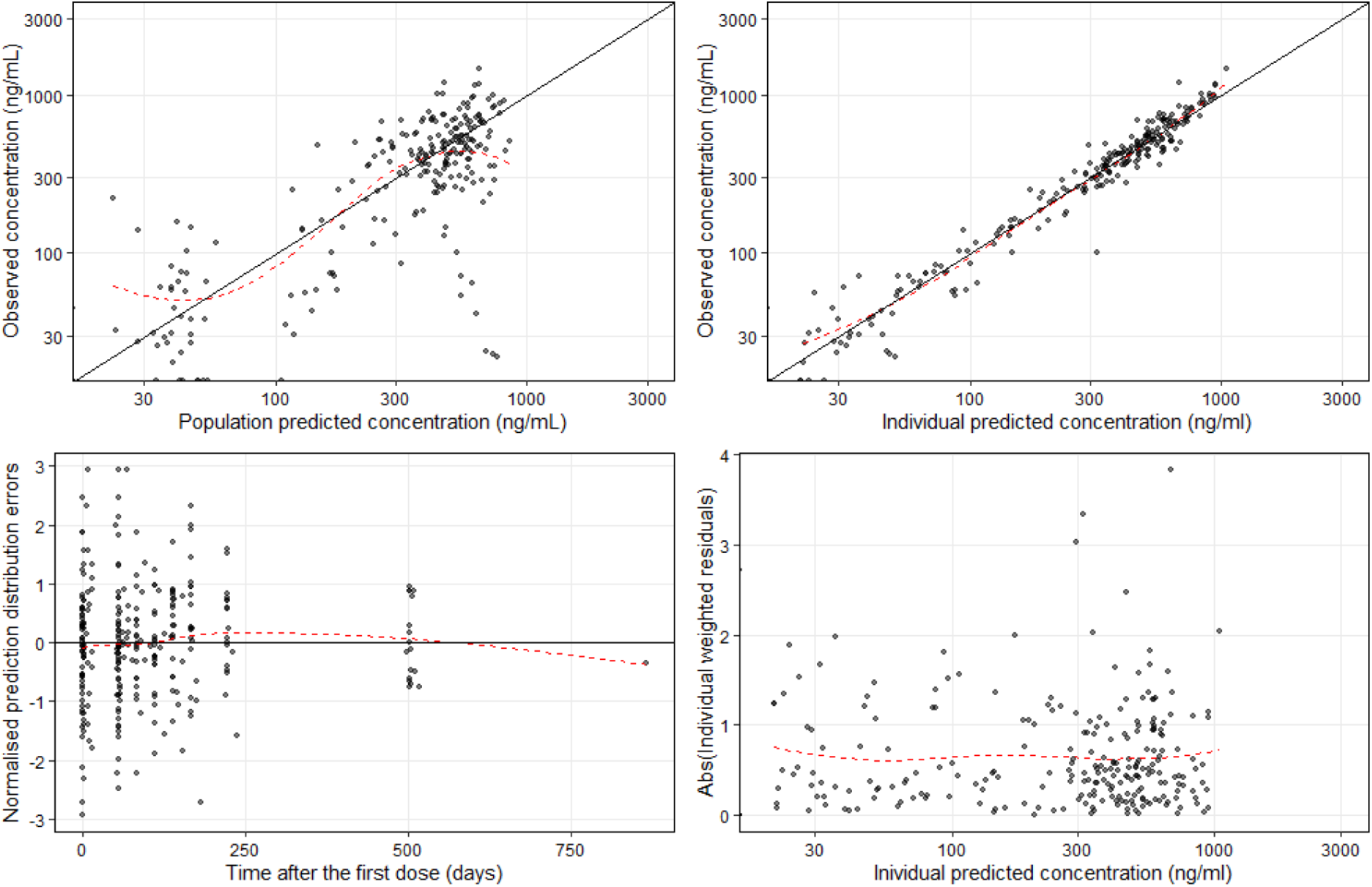
Clofazimine population pharmacokinetic model goodness of fit plots. Dots represent observations, solid black lines represent identity lines and the red dashed lines represent locally estimated scatterplot smoothing.

**Figure 2:**
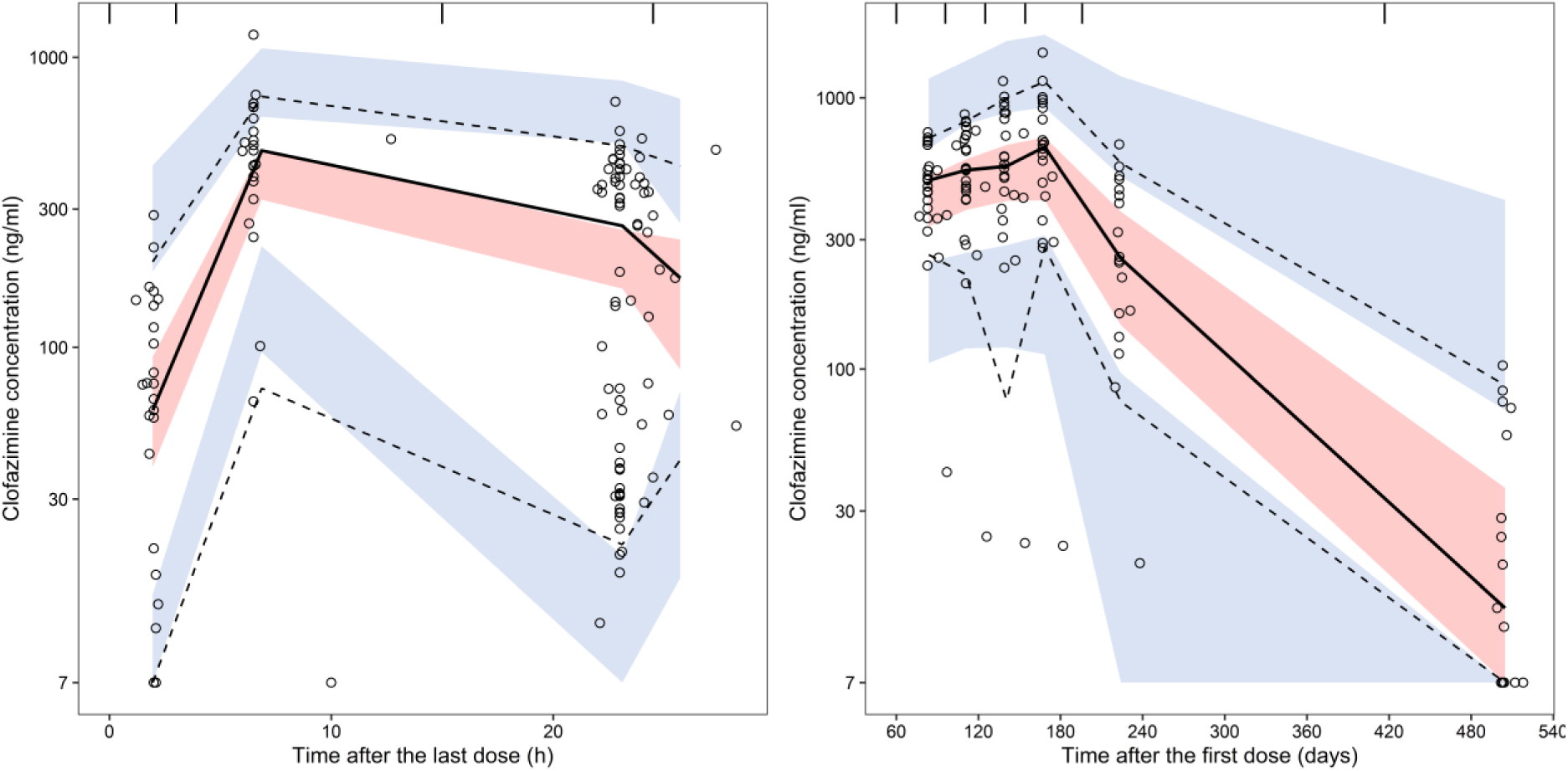
Visual predictive check, presented as time after last dose of the day 1 and week 8 visit (left panel) and time after first dose for the subsequent visits (right panel). Open circles represent the observed data, with dashed and solid lines presenting the 95% percentiles and median of the observed data. The blue and red shaded areas represent the 90% confidence intervals of model predicted 95% percentiles and median of the simulated data.

Robustness of model parameter estimates was verified using 1,000 non-parametric bootstraps. Median values of estimated parameters obtained using bootstrap analysis were consistent with corresponding final model parameter estimates, thus reflecting the final model’s robustness (Table 2). Clofazimine population PK model derived secondary parameters maximum concentration (Cmax), area under the plasma concentration-time curve (AUC), and trough concentration (Ctrough) were presented in Table 3.

**Table 3:**
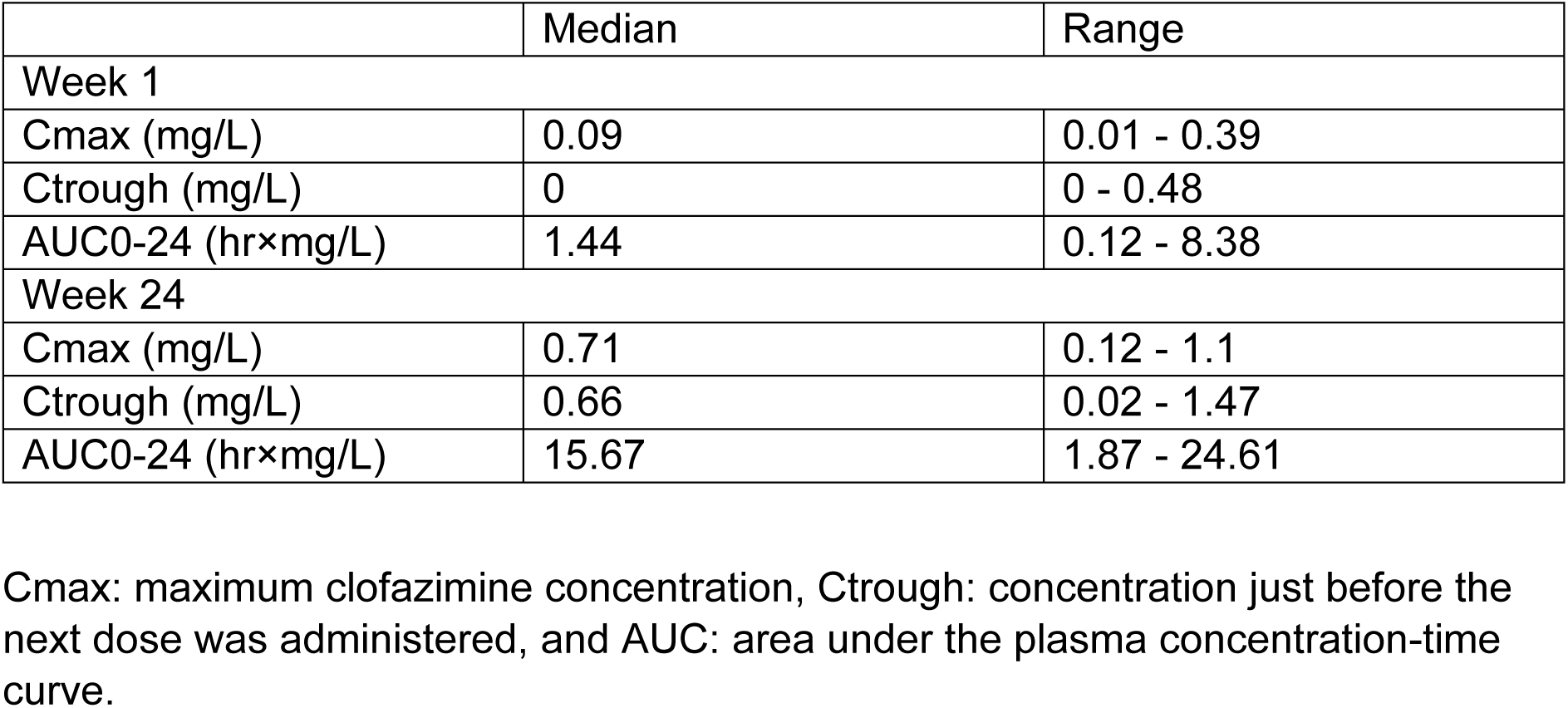
Secondary pharmacokinetic parameters derived with the clofazimine model.

The distribution of MICs of clofazimine in pure isolates of *M. tuberculosis* from 406 TB-PRACTECAL study participants disaggregated by country of enrolment are presented in figure 3. The mode MIC was 0.125mg/L and the interquartile range from 0.125 to 0.25mg/L. 99.8% of the isolates were below the interim critical concentration of 1mg/L(17).

**Figure 3:**
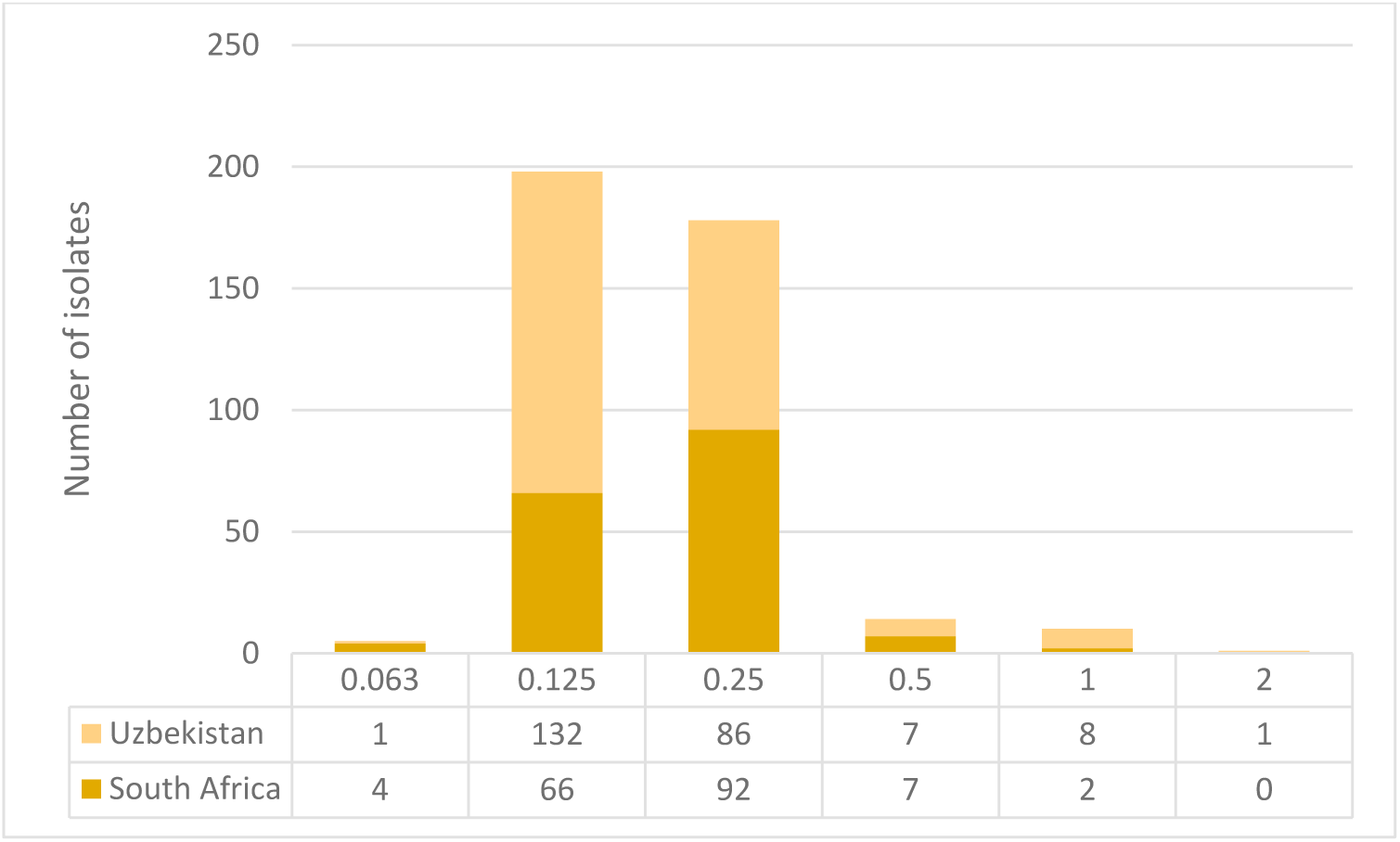
Distribution of *Mtb* baseline isolates across various clofazimine MICs (mg/L) in the TB-PRACTECAL trial.

Flat daily doses of 50mg, 100mg and 200mg, and loading dose of 200mg daily for eight weeks followed by 100mg daily were evaluated using 2,000 stochastic simulations with the observed individual patient characteristics. The PTA during the dose interval at week 24 with 0%, 85%, and 99% protein binding was evaluated.

Accounting for protein binding reduced the probability of target dramatically (Figure 4). At a dose of 100mg daily, whilst not accounting for protein binding, clofazimine plasma concentrations are maintained above 0.5 mg/L for just under 50 days but may remain above 0.032mg/L beyond 12 months (Figure 4).

**Figure 4:**
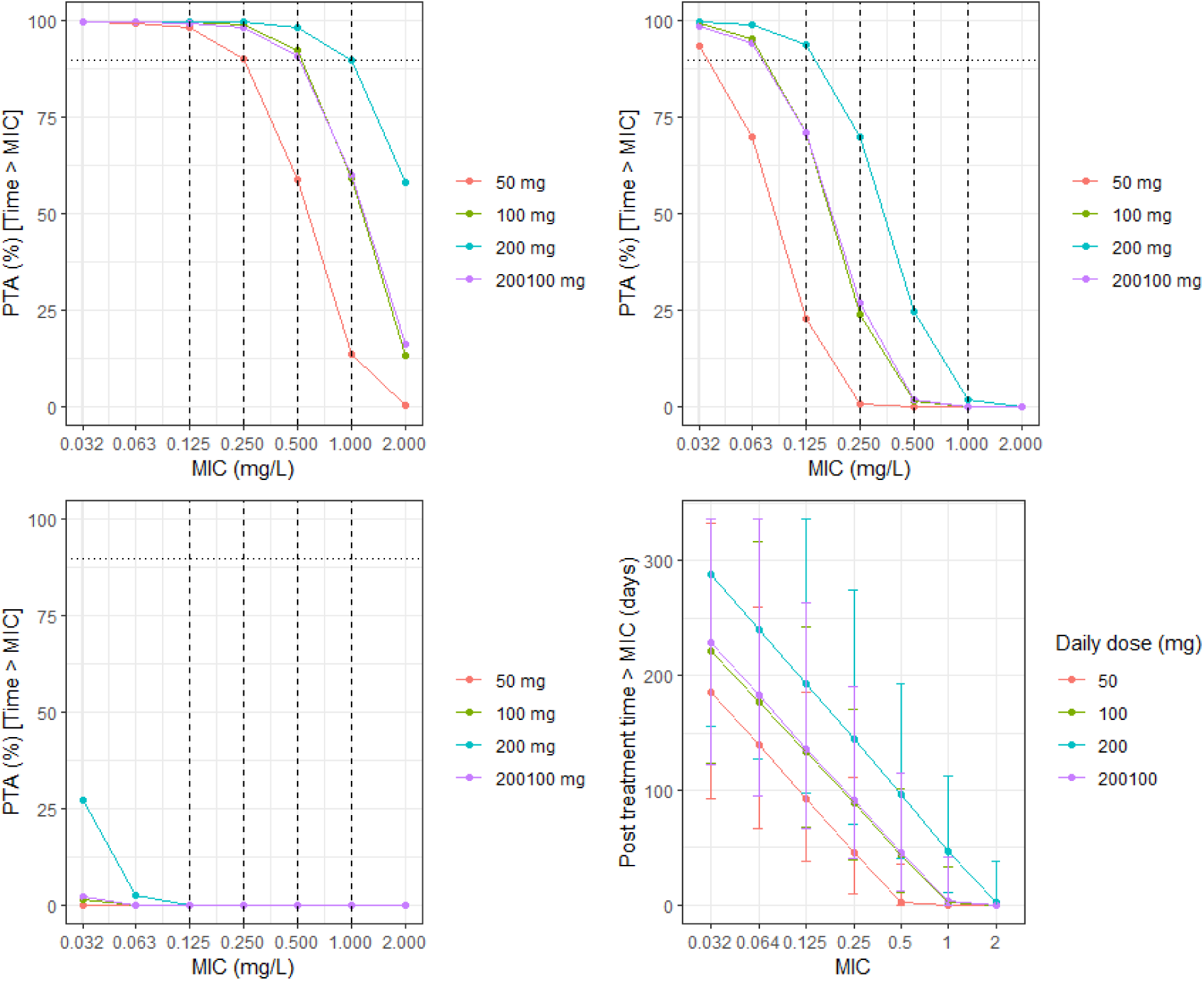
Percent Time > MIC at week 24 with 0 (top left), 85 (top right) and 99% (bottom left) protein binding scenario and time above MIC post-treatment completion assuming 0% protein binding (bottom left). 50 mg, 100 mg, and 200mg represent day dosing with the corresponding dosage whilst 200100 mg represents 200mg daily clofazimine dosing for eight weeks followed by 100mg daily for the remainder of the treatment.

## Discussion

A two-compartment first order absorption and elimination model with a lag time absorption parameter best described the PK of clofazimine in adults with rifampicin resistant tuberculosis from South Africa and Belarus. No statistically significant covariate was identified on top of weight allometric scaling.

The large volume of distribution (Vc 1,750L and Vp 9,150L) is similar to sizes reported in the other 2- and 3-compartment models (Appendix 4) (6, 8, 10, 18–20). Only one study has reported a non-body size related covariate sex, and this was attributed to differences in body fat proportion(18).

Clofazimine elimination clearance in this study was predicted at 6.84 L/hr, which is at the lower end compared to clearances reported in the adult TB patients’ studies (Appendix 4) (6, 8, 10, 18–20). Previous studies characterized the elimination phase using samples that were collected up to 14 days from the last dose which may contribute to the variability in the data for a long half-life drug. Unlike the other studies, our study on the other hand had clofazimine plasma concentration data collected at 2 and 12 months post final dose, thus closer capturing the terminal elimination phase.

Clofazimine terminal elimination half-life has been valued for treatment shortening, assuming continued anti-tubercular activity long after treatment intake has ceased(15). Moreover, clofazimine has significant activity in bacillary persister populations, and it also has 22 times higher lung and fibrous lesion concentrations compared to plasma concentrations(10, 19). Therefore, clofazimine could have a significant role post treatment prevention of recurrences. Furthermore, it may provide protection against resistance development in other anti-TB drugs with a long half-life such as bedaquiline if recurrences occur. This may explain the lower recurrences and absence of acquired bedaquiline resistant strains in the BPaLC regimen in comparison to the BPaL regimen(2).

QT prolongation and the consequent risk of Torsades du pointes have been raised as concerns when combining bedaquiline and clofazimine. However, out of 3,744 ECGs recorded over the 24 week treatment period for the investigational regimens in the TB-PRACTECAL trial, only one had a QTcF greater than 500ms(21).

Clofazimine protein binding is so high that when free drug assumptions are used in the PTA analysis, even at an MIC of 0.032 mg/L, the clofazimine concentration is almost always below the MIC (Figure 4). This may be caused by clofazimine’s likely unspecific binding to plastics in MIC experiments. A head-to-head comparison of free clofazimine PTA simulations remains impossible so long as free clofazimine is unquantified in both plasma and MIC experiments.

As a result, both the 100mg daily flat dosing and investigational 200 mg run in dose followed by 100 mg dosing, rendered a probability of the concentration remaining above the minimum inhibitory concentration above 90% at MICs of up to 0.5mg/L, not adjusting for plasma protein binding (figure 4). Within our TB-PRACTECAL study this would have meant that over 95% of the participants had *Mtb* isolates with sufficiently low enough MICs to have ≥90% T>MIC (Figure 3). Only a 200mg daily dose simulation reached the target at the 1mg/L clofazimine critical concentration.

This study has limitations, and these include a low sample size due to it being a nested sub-study. However, due to the optimal design for the timing of the samples, which was tailored for all study drugs and the 2- and 12-month post treatment completion samples, good precision and accuracy on clofazimine population PK parameter estimates was achieved(22). Moreover, parameter estimates were in line with previous reports. Clinically relevant and data driven PK-PD endpoints are lacking which complicated PTA simulations. We have used PK-PD endpoints from animal studies that reflect early bactericidal activity when clofazimine is administered as monotherapy(16). PTA simulations therefore lack translational power for informing drug combination therapy in patients.

We present a clofazimine population PK model developed using the longest post treatment follow-up data reported to date. The long follow up period adequately captures and confirms the long terminal elimination half-life of 45 days. Clinically relevant PK-PD targets need further research, especially given clofazimine’s potential niche efficacy on persister bacilli.

## Materials and Methods

Data for this study was collected as part of the PRACTECAL-PKPD sub-study, embedded within a randomized controlled trial for rifampicin-resistant tuberculosis patients(2, 22, 23). The analysis focused on the study arm where patients receive a regimen comprising daily clofazimine at 100 mg throughout the 24-week period, bedaquiline at 400 mg once daily for the initial 14 days, followed by 200 mg thrice weekly for the subsequent 22 weeks, pretomanid 200 mg daily for 24 weeks, and linezolid at 600 mg daily for 16 weeks, then reduced to 300 mg daily for the final 8 weeks.

PK blood sampling was conducted at multiple time points: Day 1 (0, 2, and 23 hours post-dose), Week 8 (pre-dose, 6.5 hours, and 23 hours), and at weeks 12, 16, 20, 24, 32, and 72 following randomization trough samples were collected. Clofazimine concentrations were measured in a certified GCP-compliant laboratory using high-performance liquid chromatography coupled with tandem mass spectrometry (HPLC-MS/MS). The quantification limit for clofazimine was established at 7 ng/mL.

R v4.1.2 was used for dataset creation, data exploration and generation of tables and plots. The list of the r packages used is in appendix 5. Clofazimine plasma concentration time series data was analysed using a population PK nonlinear mixed-effects approach with the open-source R package nlmixr2. The first-order conditional estimation with interaction (FOCE-I) algorithm in nlmixr2 was used for estimation and inter-individual variability (IIV) at the parameter level and residual variability (RV) at the observation level were used to dissect and quantify variability between patients from residual variability within a patient.

Clofazimine population PK models, available in the public domain, typically comprise multiple disposition compartments, and a one-, two-, and three-compartment model was consequently evaluated together with combined, proportional, additive and log-transformed residual error models. Linear absorption (k_a_) with and without lag-time was explored through estimation and fixing to literature values.

A correlation matrix with covariates and clearance and volume of distribution eta estimates explored associations as well as covariate collinearity. Bodyweight scaling of clearance and volume parameters included allometric scaling with power coefficients fixed to 0.75 and 1, respectively. Further covariate testing included stepwise forward inclusion (P<0.05, ΔOFV > 3.84 per degree of freedom) and backward elimination (p<0.001, ΔOFV> 10.83 per degree of freedom).

Visual inspection of model fits included Goodness-of-fit plots (GOFs) and visual predictive checks (VPCs). A non-parametric bootstrap was used to assess the robustness of the model parameter estimates and eta-shrinkage, relative standard error, and omega and sigma estimates were used to assess the precision and robustness of the model.

Minimum inhibitory concentrations were determined using MGIT and a routine testing concentration set (2, 1, 0.5, 0.25, 0.125, 0.063 mg/L), which was extended using 0.032 mg/L if required. The results from all participants from the TB-PRACTECAL trial were summarised by country of enrolment and the median and interquartile range reported.

Using 2,000 Monte-carlo simulations, using patient level characteristics from the observed participants in this study, accounted for protein binding at 0, 85 and 99%(11, 12), the clofazimine %Time>MIC at week 24 of treatment was calculated for the MIC range observed in the TB-PRACTECAL trial (0.063 – 2 mg/L) and T(days)>MIC during the 12-month post treatment period.

## Data Availability

All data produced in the present study are available upon reasonable request to the authors

## Acknowledgements

The study was funded by Medecins sans Frontieres. FK conducted the research as part of a Sir Henry Dale Fellowship jointly funded by the Wellcome Trust and the Royal Society (Grant Number 220587/Z/20/Z).

## Contributors

Conception and design: B-TN, FK, GD and DAM. Data acquisition: IM, RM, VS, SR. PK modelling: BT-N and ZH. First draft of manuscript: BT-N. Revising and approval of manuscript: all

## Ethics statement

The study was approved by the MSF Ethics Review Board (reference no. 1541) and the LSHTM Ethics Committee (reference no. 16249). The Belarus RSPCPT ethics committee and the regulator-Centre of Excellence for the Minsk site. PharmaEthics for the Don Mckenzie and Dorris Goodwin hospitals sites, University of Witwatersrand Human Research ethics committee for the Helen Joseph and King DiniZulu Hospitals sites and the South Africa Health Products Regulatory Authority

## Data access statement

Deidentified data will be available to researchers upon a written request to the Medical Director, Médecins sans Frontières, Operational Centre Amsterdam, the Netherlands.

## Funding

Médecins sans Frontières

## Appendix 1: R code for the clofazimine model

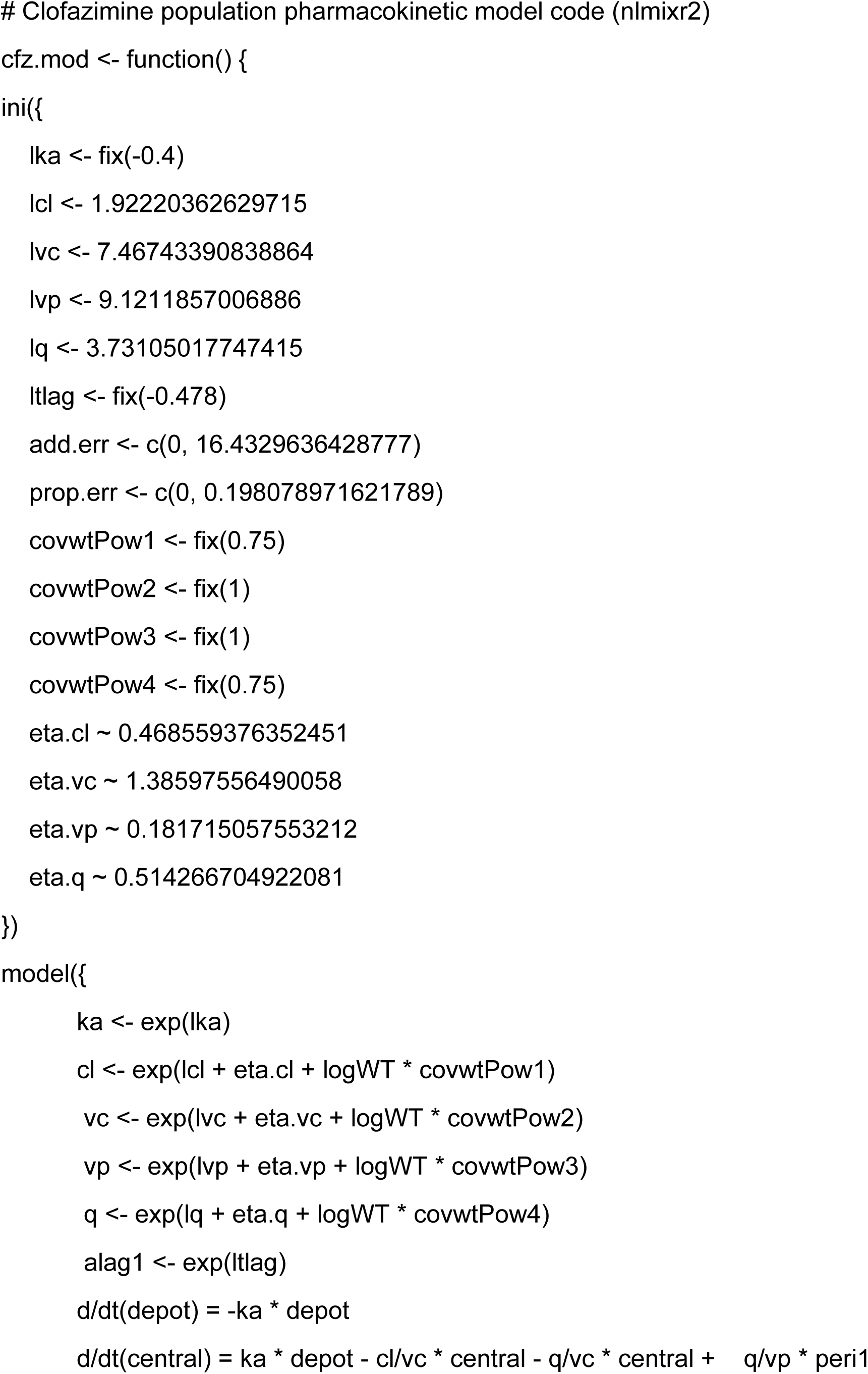
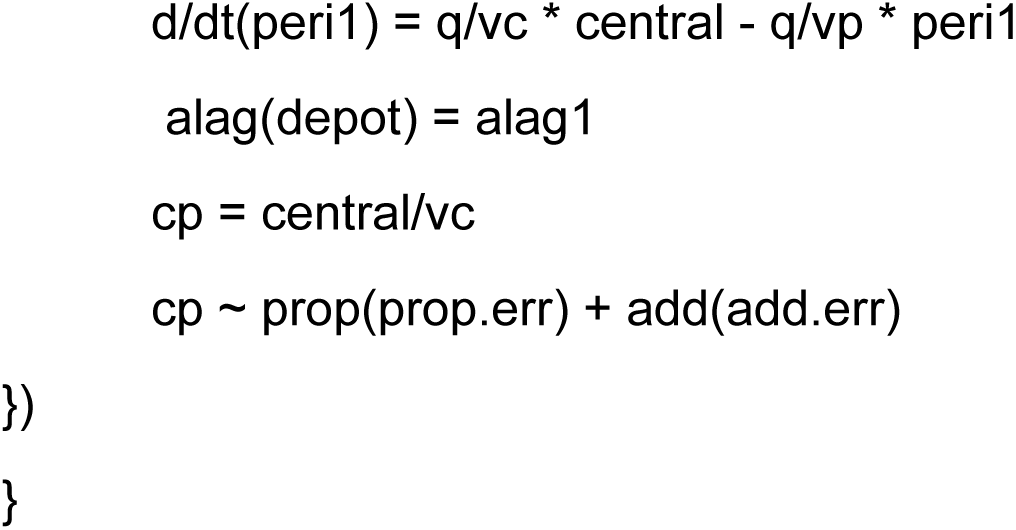

## Appendix 2: Covariate and parameter correlation matrix

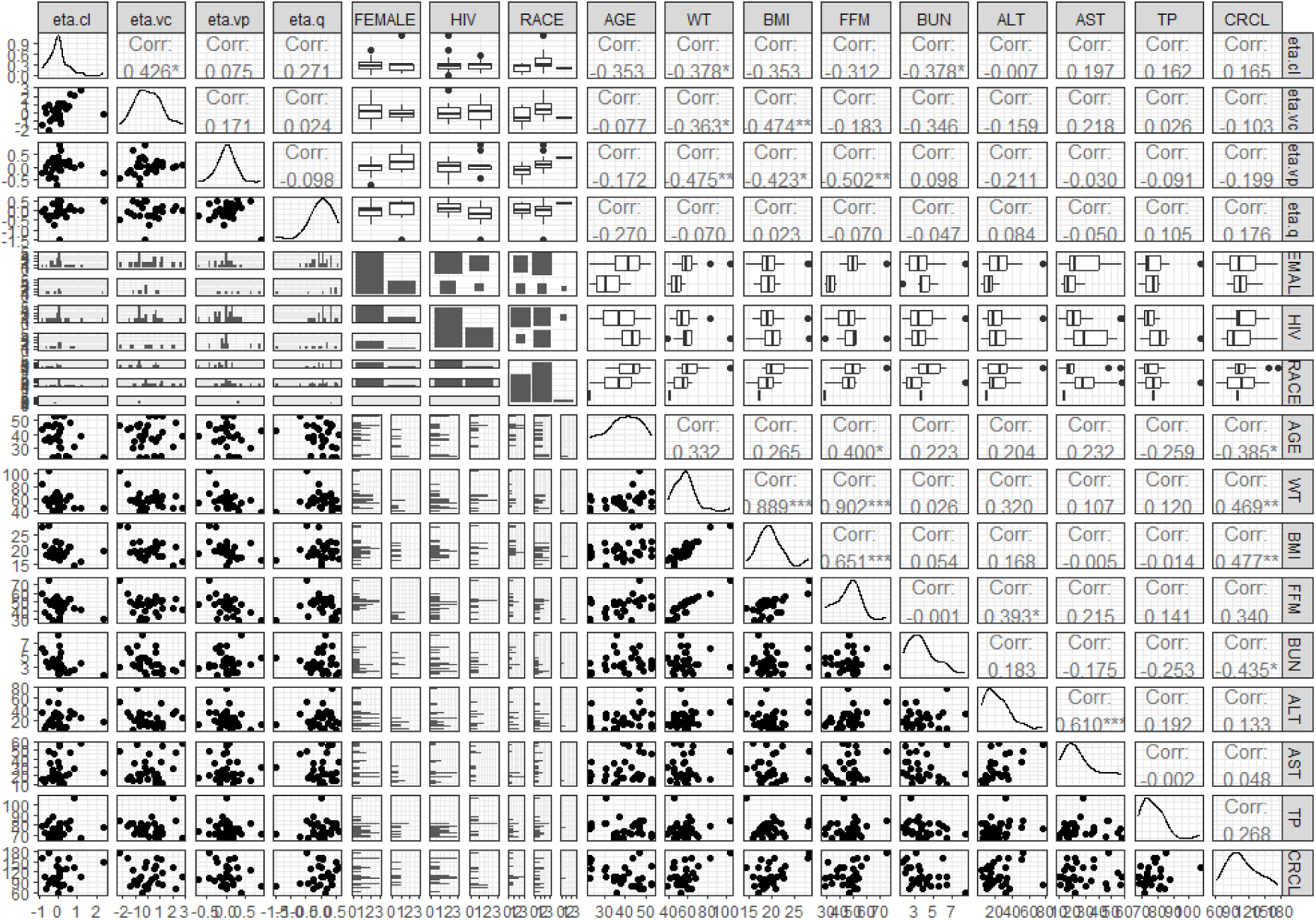

Covariate vs parameter matrix on base model. WT: weight, BMI: body mass index, FFM: fat free mass, BUN: blood urea nitrogen, ALT: alanine transaminase, and AST: aspartate aminotransferase, TP: total protein, CRCL: estimated creatinine clearance, random.g: BPaLM, BPaLC or BPaL arm.

## Appendix 3: Individual model fit plots

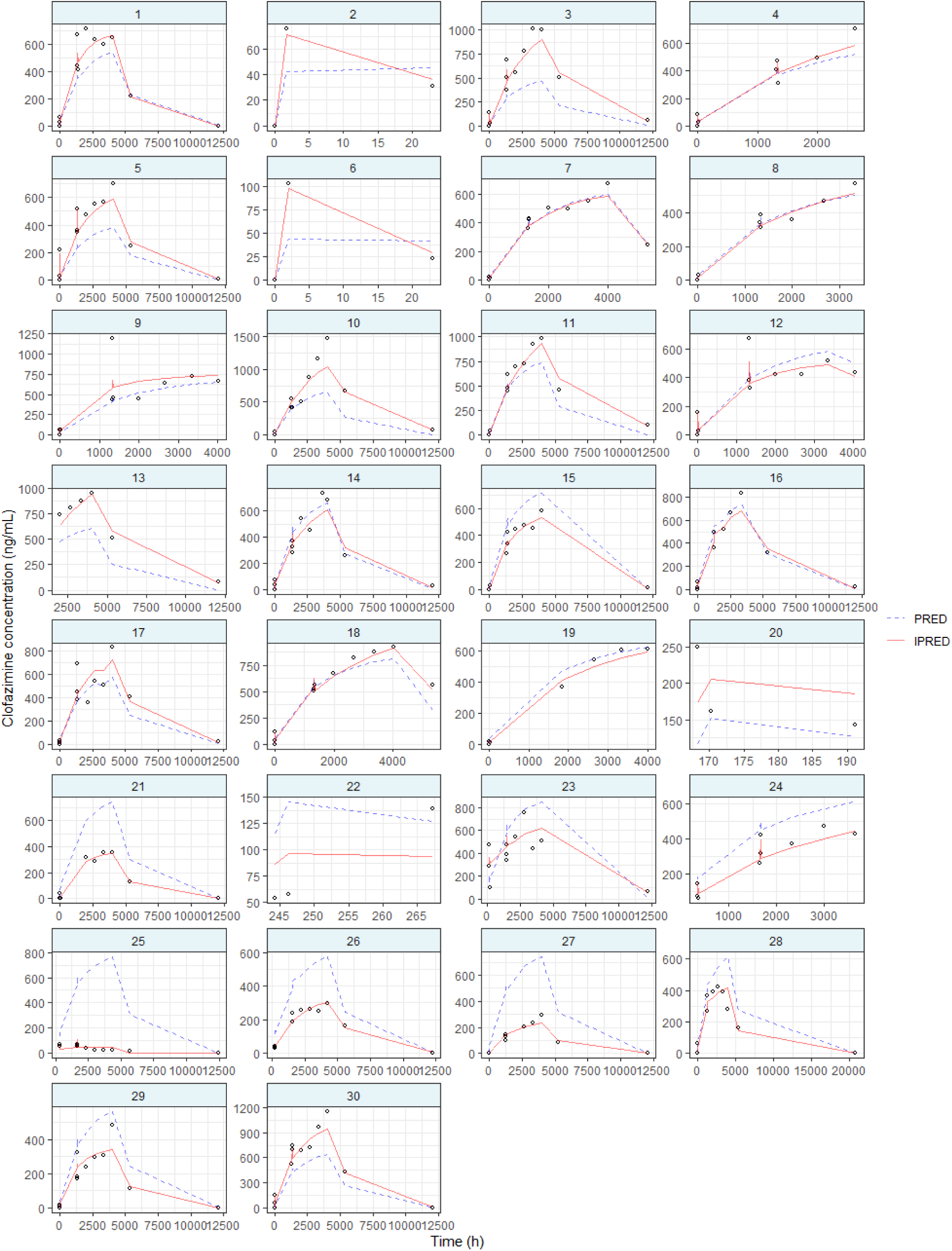

Individual linezolid plasma concentration - time profiles. Each panel represent a patient, with open circles representing observed plasma concentrations, the blue dashed line the population predictions by the developed model and the red solid lines the individual population predictions.

## Appendix 4: Previously published clofazimine population pharmacokinetic models and primary parameters

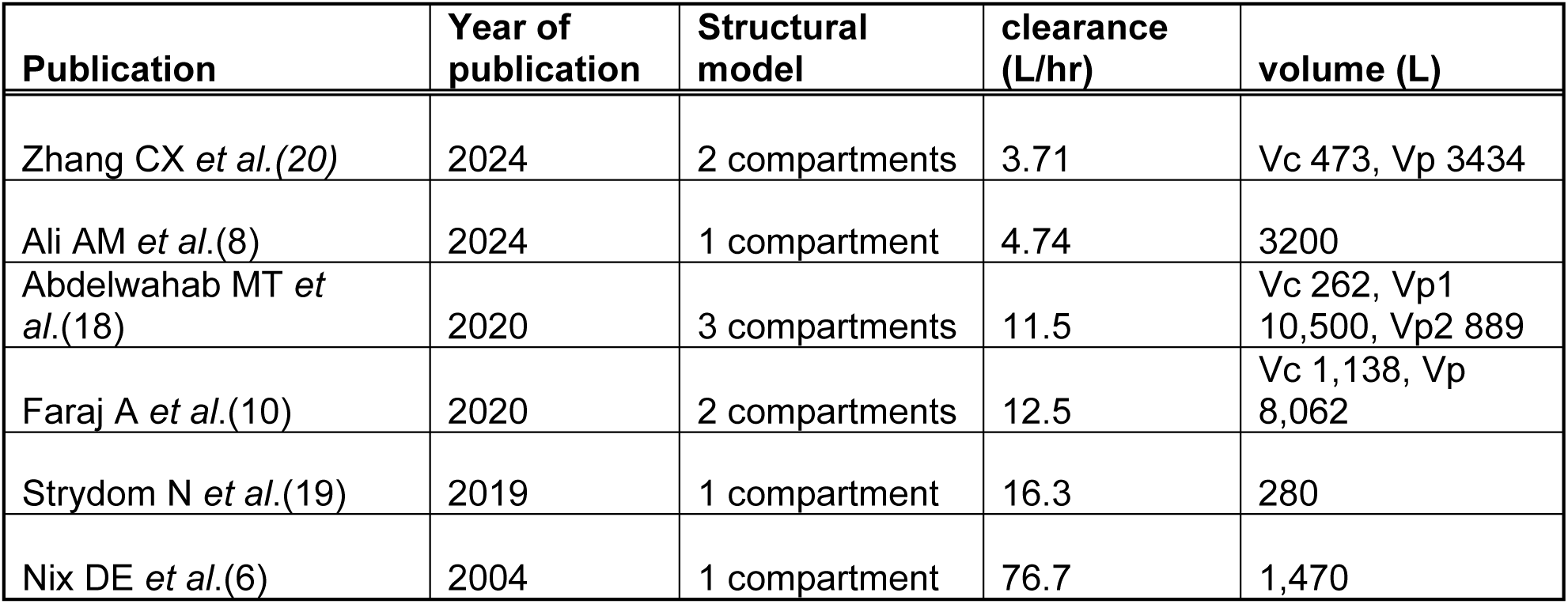

| Publication | Year of publication | Structural model | clearance (L/hr) | volume (L) |
| --- | --- | --- | --- | --- |
| Zhang CX *et al.*(20) | 2024 | 2 compartments | 3.71 | Vc 473, Vp 3434 |
| Ali AM *et al.*(8) | 2024 | 1 compartment | 4.74 | 3200 |
| Abdelwahab MT *et al.*(18) | 2020 | 3 compartments | 11.5 | Vc 262, Vp1 10,500, Vp2 889 |
| Faraj A *et al.*(10) | 2020 | 2 compartments | 12.5 | Vc 1,138, Vp 8,062 |
| Strydom N *et al.*(19) | 2019 | 1 compartment | 16.3 | 280 |
| Nix DE *et al.*(6) | 2004 | 1 compartment | 76.7 | 1,470 |

## Appendix 5: Used r packages

library(rxode2)

library(nlmixr2)

library(reshape2)

library(ggplot2)

library(tidyverse)

library(PerformanceAnalytics)

library(psych)

library(dplyr)

library(GGally)

## Notes

### Competing Interest Statement

The authors have declared no competing interest.

### Clinical Trial

ClinicalTrials.gov ID NCT04081077

### Clinical Protocols

https://doi.org/10.1136/bmjopen-2020-047185

### Funding Statement

This study was funded by Medecins sans Frontieres

### Author Declarations

The study was approved by the Medecins Sans Frontieres Ethics Review Board (reference no. 1541) and the London School of Hygiene and Tropical Medicine Ethics Committee (reference no. 16249). The Belarus Republican Scientific and Practical Centre of Pulmonology and Tuberculosis ethics committee for the Minsk site. PharmaEthics for the Don Mckenzie and Dorris Goodwin hospitals sites, University of Witwatersrand Human Research ethics committee for the Helen Joseph and King DiniZulu Hospitals sites

